# Evaluating Semantic Similarity Methods for Comparison of Text-derived Phenotype Profiles

**DOI:** 10.1101/2021.08.08.21261762

**Authors:** Luke T Slater, Sophie Russell, Silver Makepeace, Alexander Carberry, Andreas Karwath, John A Williams, Hilary Fanning, Simon Ball, Robert Hoehndorf, Georgios V Gkoutos

## Abstract

Semantic similarity is a valuable tool for analysis in biomedicine. When applied to phenotype profiles derived from clinical text, they have the capacity to enable and enhance ‘patient-like me’ analyses, automated coding, differential diagnosis, and outcome prediction, by leveraging the wealth of background knowledge provided by biomedical ontologies. While a large body of work exists exploring the use of semantic similarity for multiple tasks, including protein interaction prediction, and rare disease differential diagnosis, there is less work exploring comparison of patient phenotype profiles for clinical tasks. Moreover, there are no experimental explorations of optimal parameters or methods in the area. In this work, we develop a reproducible platform for benchmarking experimental conditions for patient phentoype similarity. Using the platform, we evaluate the task of ranking shared primary diagnosis from uncurated phenotype profiles derived from text narrative associated with admissions in MIMIC-III. In doing this, we identify and interpret the performance of a large number of semantic similarity measures for this task, and provide a basis for further research on related tasks in the area.

## Background

Analysis of natural language in clinical settings facilitates novel insights into relevant biomedical entities, which in turn lead to improved outcomes for patients [1, 2]. Biomedical ontologies are often employed as resources in NLP analyses, aiding in the resolution and reduction of ambiguity through their provision of controlled domain vocabularies and consensus definitions of biomedical concepts [3].

Moreover, linking instances of entities mentioned in text with classes in biomedical ontologies produces semantic representations of the entities described by those texts. These representations facilitate secondary analysis that make use of background knowledge encoded in or linked by ontologies. One way for achieving this is through semantic similarity: a class of methods that leverage the structural features of ontologies to calculate numerical measures of similarity between concepts, or entities described by sets of concepts [4]. These methods have been widely explored, amongst others, for prediction of protein-protein interaction [5], disease gene prioritisation [6, 7], and rare disease diagnosis [8].

While semantic similarity has been widely explored for many tasks in biomedicine, it has only more recently been applied to phenotype profiles text-mined from clinical narrative text. One work used similarity of phenotypes text-mined from literature to characterise the human diseasome [9]. Another investigation developed Doc2HPO, which uses a hybrid approach of concept recognition with expert curation for patient phenotyping, then passing those phenotype profiles to variant prioritisation programs [10]. Another work explored the use of uncurated text-derived phenotypes for differential diagnosis of common diseases [11].

Benchmarking tasks have been defined for the comparison of semantic similarity measures in several biomedical spaces. For example, the CESSM task supports collaborative evaluation of semantic similarity measures for tasks involving the Gene Ontology(GO) [12]. In the clinical space, experiments were performed to evaluate effectiveness at measuring similarity between classes in medical terminologies [13, 14]. However, there are no such evaluations for tasks pertaining to predictive clinical tasks using patient phenotype profiles, despite this being an emerging area of application for semantic similarity. Nor are there any evaluations for such tasks made using text-derived phenotype profiles.

Previous work comparing semantic similarity methods on a single hierarchical medical terminology, SNOMED-CT, revealed poor agreement between methods [14]. Other works comparing semantic similarity methods for different tasks using GO found that different methods were more suitable for different tasks [4]. Differences may also arise from alternative design principles and structures of the ontology being used to calculate similarity scores.

Furthermore, the use of semantic similarity on large text-derived phenotype profiles is challenging. In one recent work, concept recognition performed on clinical narrative associated with 1,000 MIMIC-III admissions with the Human Phenotype Ontology (HPO) [15] producing 43,953 separate annotations, with a mean of nearly 44 annotations per admission [16]. Other work has shown that greater annotation size leads to bias in semantic similarity calculations, in the form of greater similarity between entities with more annotations, regardless of whether they are more biologically related [17]. These findings reveal the need for investigations comparing semantic similarity methods for clinical tasks using phenotype profiles, and for clinical tasks using uncurated text-derived phenotype profiles.

In this work, we describe the development of a platform for reproducible and repeatable evaluation of different experimental conditions for comparison of patient phenotype profiles. We use this platform to compare and report upon performance of different semantic similarity methods for predicting shared primary patient diagnosis using uncurated text-derived phenotype profiles, and report upon the results of this investigation, identifying best-performing methodological configurations.

## Methods

We developed a framework with which to perform reproducible semantic similarity experiments using HPO-based phenotype profiles, using the MIMIC-III dataset [18]. While the software can be used to perform experiments with phenotype profiles of any etiology, for the purposes of our investigation we developed a workflow that creates uncurated text-derived phenotype profiles for evaluation. The modular nature of the implementation allows for alternative means of producing or importing phenotype profiles. The platform is implemented in the form of Jupyter Notebooks containing executable code for repeating, modifying, and evaluating the experiments. The software is available, with instructions, from https://github.com/reality/mimpred. The overall experimental methodology, visualised in Figure 1, centres around sampling admissions, obtaining the associated clinical narrative text, producing phenotype profiles, then calculating and evaluating a large set of semantic similarity methods.

**Figure 1.**
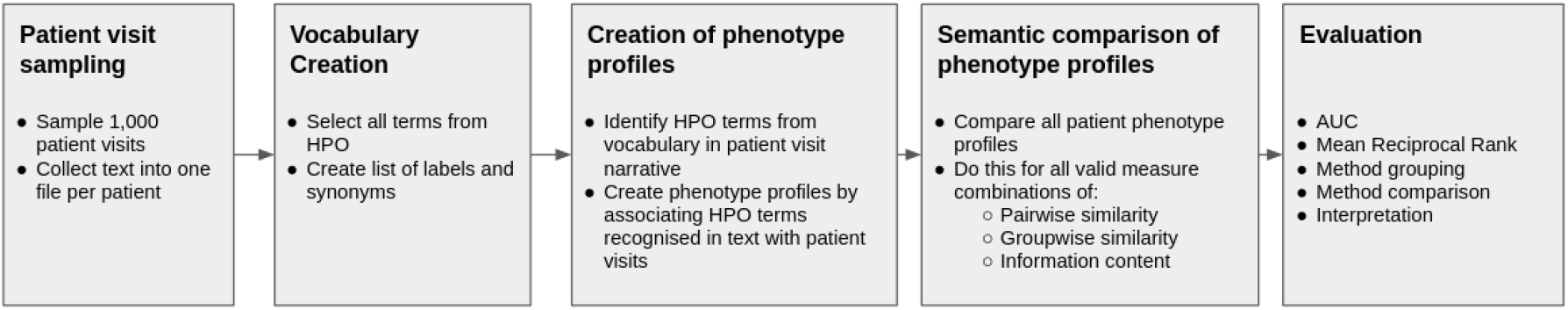
Overall description of the experimental methodology.

### Sampling Admissions

Our evalution employed the MIMIC-III database [18]. MIMIC is a freely available database of healthcare data describing nearly 60,000 visits to a critical care clinic at Beth Israel Deaconess Medical Center in Boston, Massachusetts. It provides a wealth of structured and unstructured data concerning those visits, including clinical narrative text. Diagnoses for patients are also provided in the form of ICD-9 codes, which were produced following visits by professional curators.

We sampled 1,000 admissions, each describing a single patient visit, from the MIMIC dataset, collecting their associated texts together into one file per patient visit. We also performed pre-processing to collapse multiple line breaks into sentence breaks, and otherwise to improve formatting of the files. Finally, we associated each admission with its primary diagnosis given by structured coding data.

### Vocabulary Creation and Creation of Phenotype Profiles

We then used the Komenti semantic text mining framework [19] to create a vocabulary from all non-obsolete terms in the Human Phenotype Ontology (HPO), which describes phenotypic abnormalities in humans [15]. Komenti was then applied annotate the texts, identifying HPO terms from the vocabulary in the clinical narrative associated with each admission. The set of HPO terms identified with each admission constitutes that the phenotype profile for that admission.

### Semantic Comparison of Phenotype Profiles and Evaluation

Subsequently, we created sets of similarity scores for each pairwise combination of phenotype profiles derived from the text associated with the 1,000 sampled admissions. To calculate the semantic similarity scores, we used the Semantic Measures Library toolkit[20], and we explored every available combination of information content, pairwise, and groupwise similarity measure available within the library.

Pairwise similarity methods calculate similarity between a single pair of terms in a biomedical ontology. These can be structural methods, which use some heuristic derived from the structure of the ontology when considered as a graph, such as through simple distance in the ontology graph, or distance to a common ancestor. Pairwise similarity methods can also be defined in terms of an information content measure. An information content measure is a method that determines how specific or informative a particular term is. These can also be structural, such as in the case of the Zhou [21] method, which uses a measure of how deep a term is in the ontology graph, or corpus-based, such as the Resnik [22] method, which uses a measure of how likely a term is to appear in a corpus. Each pairwise similarity method that admits an information content measure will use it in a different way, such as being defined as the information content of the greatest common ancestor of the two input classes.

Phenotype profiles, however, generally contain more than one phenotype to describe an entity. To calculate semantic similarity between two sets of ontology classes, groupwise measures can be employed. These are further delineated into two groups. Direct groupwise measures define their own methods of directly calculating similarity between two sets of classes. These are distinguished from indirect groupwise measures, which admit a pairwise similarity method, and employ this as a constituent of the method. For example, the max indirect groupwise measure compares each term in the two input sets using a given pairwise similarity method, and then selects the greatest score as the result.

Since there are distinctions between pairwise methods that may or may not admit information content measures, and groupwise measures that may or may not admit pairwise similarity measures, we had to identify the full set of combinations of available methods. In particular, we evaluated combinations of every indirect groupwise measure and every pairwise measure, as well as an additional combination of each IC-using pairwise measure with each IC measure. This led to 300 total experimental settings. We encoded these configurations into a set of XML files that can be used as parameters for the Semantic Measures Toolkit.

Each experimental setting, defining a method of measuring similarity between phenotype profiles, produces a pairwise similarity matrix when used to compare every phenotype profile with every other phenotype profile. Subsequently, each of these was was transformed into a set of vectors that associated the semantic similarity score for each pair of admissions with the outcome: whether those admissions shared a primary coded diagnosis. This process is described in Figure 2

**Figure 2.**
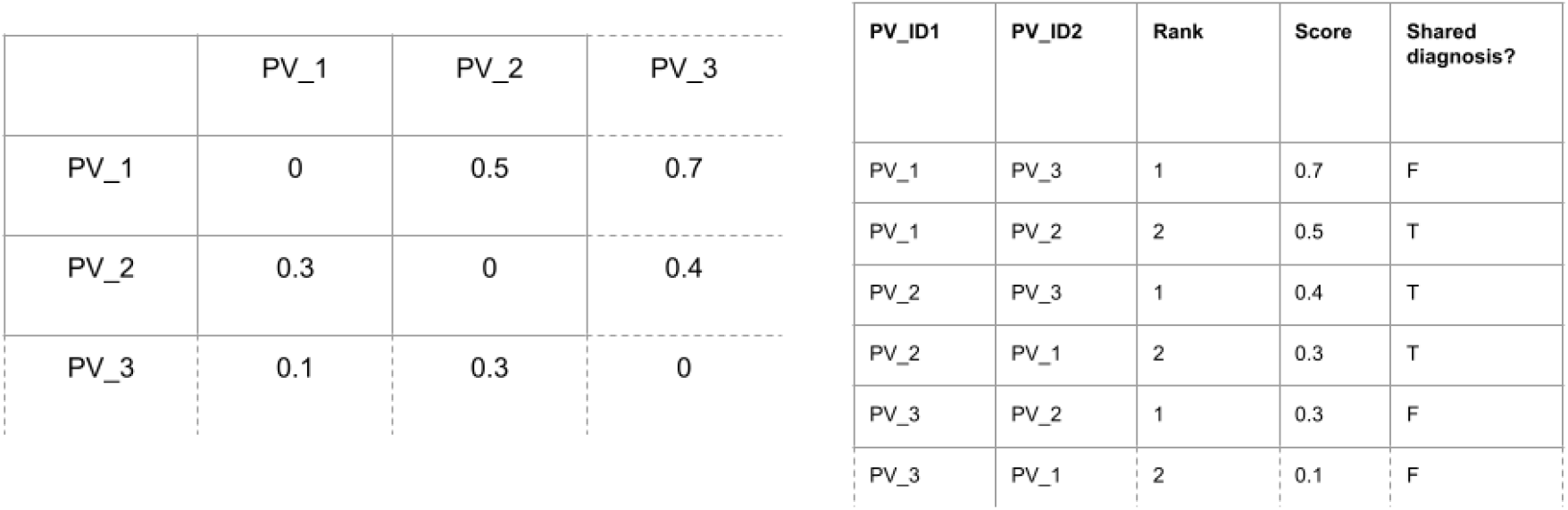
A sample of an example output of the semantic similarity process. On the left is a semantic similarity matrix, in which every phenotype profile associated with a patient visit has been compared with every other one. The result is a matrix of similarity values. To evaluate the matrices, we then convert these into a ranked list of similarity values for each patient visit, which also includes whether or not the two patient visits being compared share a primary diagnosis. The latter structure is used to create our evaluation scores (e.g. AUC). In the experiment described in this article, this process is repeated once for every combination of semantic similarity measure being explored, since each will produce a separate similarity matrix.

We used the association between pairwise admission scores and shared diagnosis to evaluate how predictive those scores were of shared primary admission diagnosis. We evaluated performance using Area Under the receiver operating characteristic Curve (AUC), Mean Reciprocal Rank (MRR), and Top Ten Accuracy (the percentage of patients for whom the correct diagnosis was in the top ten most similar entities). One previously identified limitation of analyses that compared patient phenotype profiles for prediction of shared diagnosis was that no correct diagnosis can be found for an admission if there is no other admission in the set with a matching primary diagnosis [23]. To identify whether, and to what extent, a lack of matching diagnoses for any admissions in our sample negatively affected scores, we also evaluated a modified MRR metric that removed values of MRR that were 0, since values of 0 indicate there were no admissions with a matching primary diagnosis (and therefore the correct label did not appear at all in the ranking).

## Results

We created phenotype profiles for 1,000 admissions from MIMIC-III, by identifying HPO terms in their clinical text narrative. We then created semantic similarity matrices for these phenotype profiles using each available combination of pairwise, groupwise, and information content measure. A break-down of different SS and IC method categories is given in Table 1. With the loss of 60 combinations due to two non-functioning pairwise measures, we obtained 300 results. Using these similarity matrices, we evaluated how predictive the semantic similarity score was of shared primary diagnosis.

**Table 1.**
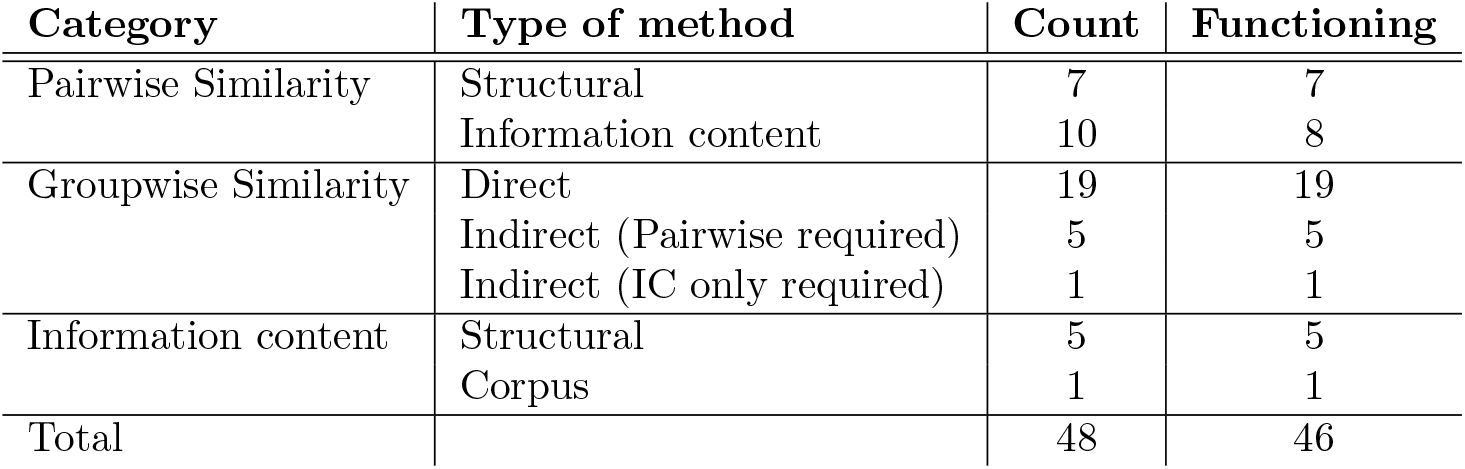
Breakdown of the different categories of semantic similarity measures available in the Semantic Measures Toolkit.

| Category | Type of method | Count | Functioning |
| --- | --- | --- | --- |
| Pairwise Similarity | Structural | 7 | 7 |
|  | Information content | 10 | 8 |
| Groupwise Similarity | Direct | 19 | 19 |
|  | Indirect (Pairwise required) | 5 | 5 |
|  | Indirect (IC only required) | 1 | 1 |
| Information content | Structural | 5 | 5 |
|  | Corpus | 1 | 1 |
| Total |  | 48 | 46 |

Figure 3 shows the distribution of scores for each evaluation metric. In the case of AUC, a majority of algorithms fall between 0.5 and 0.6, indicating performance approaching that of a random classifier, and thus that there is no signal in these methods. Interestingly, all other metrics show a median grouping of scores above what would be expected for a completely random classifier. This indicates that measures which performed poorly by AUC, may nevertheless produce at least one highly ranked correct match, despite overall scoring leading to very poor performance.

**Figure 3.**
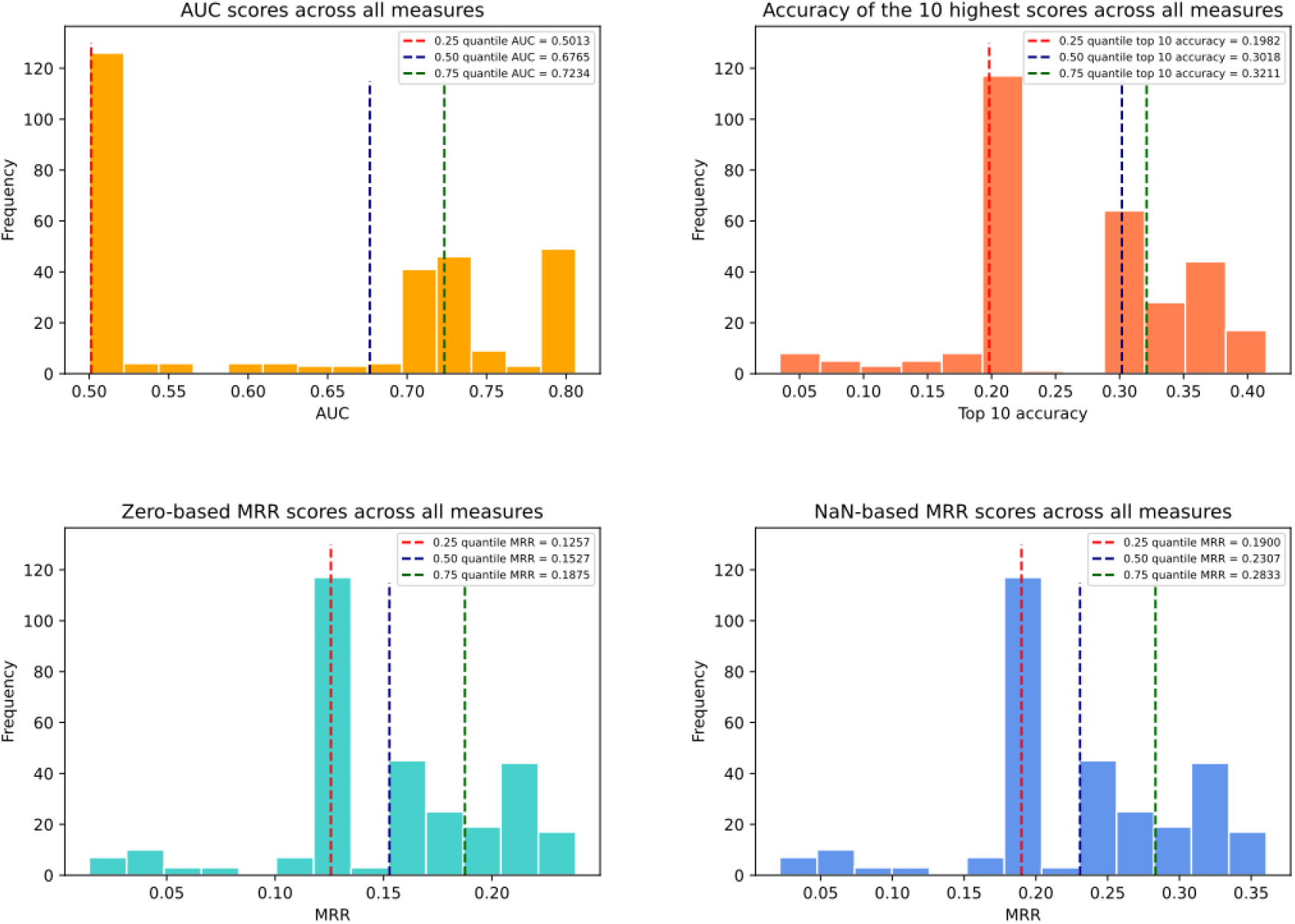
Distribution of scores using different measures. The distribution of MRR-0 and MRR-NA are the same, since these scores have a static relationship.

Nevertheless, ranking of measures correlated mostly very strongly between evaluation measures, which we characterised using Pearson correlation. Between AUC and A@10 scores this was 0.88, while between AUC and MRR it was 0.84, and MRR with A@10 was 0.97. Despite this very strong correlation, there are significant exceptions, particularly at the top end of the scores. Table 2, which lists the top three settings for each evaluation measure, confirms this. The GIC (Graph Information Content) measure [24], which admits an information content measure, accounts for all three top measures by A@10, and the top two measures by MRR. The particular information content measure makes minimal contribution to differences in performance here, with all settings being nearly identical by all evaluation measures. However, the AUC of all settings involving GIC are substantially lower, at an average of 0.67, than the best performing setting by AUC, which have an average of 0.81. The two GIC settings that lead the MRR rankings also have AUC values much lower than the Bader method that appear in third, which otherwise has nearly identical metrics, excepting a slightly reduced A@10 value. Inversely, however, the best performing measures by AUC suffer from reduced values of the other metrics, which can lead to substantially different results in a practical sense; for example, the best performing measure by A@10 has a matching patient diagnosis in the top ten in 8% more cases than does the best setting by AUC.

**Table 2.** Top algorithms by AUC, MRR, and A@10. Since MRR-NA and MRR-0 are statically dependent, the top algorithms for both are equivalent, and so only ‘MRR’ is listed here.

| By | Method | AUC | MRR-NA | MRR-0 | A@10 |
| --- | --- | --- | --- | --- | --- |
| AUC | AVG (GW) + Resnik (PW) + Resnik (IC) | 0.81 | 0.29 | 0.19 | 0.34 |
|  | AVG (GW) + Jaccard (PW) + Zhou (IC) | 0.8 | 0.28 | 0.19 | 0.32 |
|  | AVG (GW) + NODE_SIM (PW) + Zhou (IC) | 0.8 | 0.27 | 0.18 | 0.31 |
| MRR | GIC (DGW) + Zhou (IC) | 0.68 | 0.36 | 0.24 | 0.41 |
|  | GIC (DGW) + Seco (IC) | 0.68 | 0.36 | 0.24 | 0.41 |
|  | Bader (DGW) | 0.77 | 0.36 | 0.24 | 0.4 |
| A@10 | GIC (DGW) + Resnik (IC) | 0.68 | 0.36 | 0.24 | 0.42 |
|  | GIC (DGW) + Sanchez (IC) | 0.67 | 0.36 | 0.24 | 0.42 |
|  | GIC (DGW) + Min (IC) | 0.67 | 0.35 | 0.23 | 0.42 |

All three best performing methods by AUC used the AVG method of indirect groupwise comparison, which averages the result of comparisons made with its associated pairwise method. This could reflect the greater ability of the average method to encode information from large sets of annotation profiles. This is confirmed by the average metric values shown in Table 3 where the best measure by AUC was the AVG, although MRR and A@10 found higher average values through direct measures, which is also in accord with the top-scoring methods shown in Table 2.

**Table 3.** Average performance per groupwise measure.

| Groupwise Method | AUC | MRR-NA | MRR-0 | A@10 |
| --- | --- | --- | --- | --- |
| MAX | 0.51 | 0.19 | 0.13 | 0.22 |
| MIN | 0.50 | 0.18 | 0.12 | 0.19 |
| BMA | 0.67 | 0.29 | 0.19 | 0.33 |
| BMM | 0.66 | 0.22 | 0.14 | 0.28 |
| AVG | **0.7** | 0.24 | 0.16 | 0.28 |
| Direct | 0.68 | **0.31** | **0.20** | **0.36** |

Table 4 shows average scores for each information content measures, compared with the average scores for methods that did not admit IC methods. This shows that non-IC methods performed better on average by all methods, and with clear separation in all cases. Despite this, we can see another example of deviation from average trends at the top end of performance by noting that the best-performing methods by all three metrics used information content measures.

**Table 4.** Average performance from different IC measures

| IC Method | AUC | MRR-NA | MRR-0 | A@10 |
| --- | --- | --- | --- | --- |
| Resnik | 0.6 | 0.23 | 0.15 | 0.26 |
| Zhou | 0.60 | 0.22 | 0.14 | 0.25 |
| Seco | 0.60 | 0.22 | 0.14 | 0.25 |
| Sanchez | 0.6 | 0.22 | 0.14 | 0.25 |
| Max | 0.6 | 0.22 | 0.14 | 0.25 |
| Min | 0.61 | 0.22 | 0.15 | 0.26 |
| Non-IC | **0.66** | **0.27** | **0.18** | **0.32** |

## Discussion

Our experiments revealed a wide range of performances associated with combinations of methods for similarity-based prediction of shared primary diagnosis from text-derived phenotypes, measuring between very poor and good.

Despite an overall strong correlation between performance measures, the best-performing methods differed depending on the evaluation metric. This can be accounted for by considering what the metrics measure. MRR measures, for each admission, the average position of the first matching admission, A@10 measures the percentage of admissions with a matching diagnosis in the top ten most similar admissions, and AUC measures the overall ranking of all matching pairs of admissions, and their position relative to non-matching pairs. Indirect groupwise measures favoured AUC, implying that these produce better overall rankings of shared diagnoses, while direct groupwise measures favoured MRR and A@10, showing that these methods are better for producing highly ranked first matches. These differences have implications for any practical uses of similarity-based methods in a clinical environment, depending on the manner in which they might be implemented. Methods performing better on MRR and A@10 metrics may be better for direct use by humans, since these are more likely to produce highly ranked correct matches. On the other hand, hybrid approaches that make secondary use of for computation of diagnosis may benefit from measures that perform better by AUC, since aggregated information from multiple matches may be synthesised for improved prediction. Moreover, the AUC measure is derived from similarity score rankings that are calculated globally for all admission pairs, rather than per-admission (as A@10 and MRR are). This means that AUC considers similarity score rankings in global context. As such, it takes into account that while incorrect pairs may be highly ranked in the context of a single admission, the overall ranking of the pair may be appropriately low. This helps to measure the negative predictive power of the approach through its ability to express a low score, where a good match cannot be found. Use of this information in a practical setting would require further development, however, such as the addition of a likelihood ratio or percentage based on a suggestion’s global ranking.

By AUC, settings involving the AVG indirect groupwise measure performed best, while the other two metrics favoured GIC. These methods are in fact relatively similar, with GIC being defined by the average information content of the intersection of terms in the considered sets [24], and AVG being defined by the average result of pairwise comparisons between all terms in the two sets. The major difference between these methods is that GIC considers only exact term matches between the two sets of terms, while AVG admits pairwise methods that measure similarity between individual terms, such as through their most informative common ancestor, and not requiring an exact match (which is the case for the best performing case, using Resnik). This could account for the difference in performance measured by different metrics, as admission pairs with many exact matches would be very highly ranked, while admissions with similar, but not exactly matching, phenotypes, will be lowly ranked in favour of pairs with exactly matching but irrelevant phenotypes (for example, those which are common in the corpus, such as ‘pain’). As such, GIC may produce highly ranked correct answers from pairs where there are an abundance of exactly matching phenotypes, but suffer in cases with fewer exact matches, leading to a lower AUC score. This is an interesting result, that highlights the respective benefits of alternative methodological choices. It is possible that further investigation could be undertaken to identify whether a synthesis of exact and similarity-based methods of semantic similarity could be used to create an approach that combines their virtues. Perhaps, making use of both a local ranking and global ranking, derived from different similarity methods, could lead to a superior method.

The finding that AVG was the best performing indirect groupwise measure by AUC conflicts somewhat with findings for applications to other problems, particularly in relation to genetics and variant prediction [4]. Despite average values for performance being greater for non-IC measures, the best performing measures overwhelmingly involved IC measures, which is in accord with other findings and applications. By intuition, experiments involving uncurated text-derived phenotypes should benefit from IC measures, since these will downregulate annotations that are erroneous or uninformative. It is perhaps surprising that structural methods performed almost exactly equivalent to the Resnik annotation frequency method, showing that structural specificity of a term, rather than its probability of appearing in a corpus, is a better measure of information in this case. This implies that error from these profiles derives more from an abundance of over-general ontology terms, rather than overall abundance of terms made by erroneous annotation or commonality in the source corpus.

The significance of the difference between MRR-NA and MRR-0 values is that the MRR-NA value shows us what the MRR value would be if every patient had a match. That these values are much greater, implies that the MRR values are being negative impacted by admissions without a matching admission (one that shared a primary diagnosis). This, in turn, implies that a larger sample size of patients may improve the MRR-0 value close to the MRR-NA value, since there would be fewer unmatched admissions.

We also tested the setting that was used in our previous work, that is the combination of Resnik pairwise, BMA, and Resnik IC [23]. In this experiment, this yielded an AUC of 0.73, and an MRR-0 of 0.23, lower than the previously reported results of 0.77 and 0.42, respectively. We believe this to be caused by patient selection: in the previous experiment, admissions were limited to those with a primary diagnosis that contained a direct mapping in the Disease Ontology (DO), to facilitate exploration of differential diagnosis with DO disease profiles. As such, we can conclude that the results reported in that paper are optimistic with respect to the full set of diseases described in MIMIC. In addition, these results show that improved performance may have been obtained in this experiment by using AVG as the indirect groupwise measure, instead of BMA.

Another contribution of this work is the experimental platform, which makes it easier for researchers to investigate other experimental settings for this kind of problem. It would be possible to introduce different methods of producing text-derived phenotype profiles, curating those profiles, or evaluating prediction of different outcomes. While our investigation compares ability to classify primary diagnosis, which we use as an indicator of success in measuring true semantic similarity, it does not go further to evaluate grouping on additional factors our outcomes. These tasks require further investigation and development of tailored methods. We hope that the experimental platform described in this work will form the basis for evaluation studies, in a manner similar to those previously described for tasks involving genes and gene products [4].

One such area that the experimental platform could be turned towards, is the prediction of patient diagnosis through comparison with disease profiles, rather than with other patients. In our previous work, we identified that prediction of patient diagnosis from text-derived phenotypes was best when using SS to compare patients to disease profiles mined from literature, and extended with in-context training from patient profiles [23]. We plan to follow this study up with another, comparing patients with different sizes and derivations of disease profile. We do not expect the results of this analysis to necessarily recapitulate the results described in this article, due to the asymmetry of entity types being compared, along with differences in annotation size, source, and composition.

Extension of this work should also consist in its application to different clinical settings. MIMIC-III describes admissions to a critical care setting, which holds particular biases with respect to common diseases and treatment, as well as towards the particular hospital from which the data is derived. As such, results and even best-performing methods may vary depending on many related factors, such as geographic location, jurisdiction, department, or clinical focus.

While this work tests all available methods implemented in the semantic measures library, there are also many semantic similarity approaches implemented as R packages [25], web-based tools [26], and standalone software [27]. It is possible that these approaches could hold the potential to be applicable to the problems considered here, and this is an area for future exploration. Many of these tools, however, focus exclusively on basic biology, and Gene Ontology in particular, and so additional adaptation may be required to fit them to the problem domain.

## Conclusion

We have presented the development of a platform for evaluating semantic similarity methods for tasks using patient phenotype profiles. We used this implementation to evaluate a large number of settings for the task of predicting shared primary diagnosis from uncurated text-derived phenotype profiles. We interpreted a large range of results by multiple measures, and identified methods that performed more optimally. These results, along with the platform, help to provide a basis for systematically identifying and evaluating methods for practical clinical tasks using semantic similarity methods.

## Data Availability

Software is available from the link. MIMIC data must be acquired separately according to the relevant guidelines.

https://github.com/reality/mimpred

## Ethical approval

This work makes use of the MIMIC-III dataset, which was approved for construction, de-identification, and sharing by the BIDMC and MIT institutional review boards (IRBs). Further details on MIMIC-III ethics are available from its original publication (DOI:10.1038/sdata.2016.35). Work was undertaken in accordance with the MIMIC-III guidelines.

## Availability of data and material

The framework developed to run this experiment is freely available from https://github.com/reality/mimpred. The annotated MIMIC dataset is not made publicly available, because researchers are required to meet ethical conditions to access MIMIC-derived datasets. However, this can be individually applied for and downloaded separately for use with our software.

## Acknowledgements

We would like to thank Dr Paul Schofield for helpful discussions and use of server resources.

